# One-year results of long femoropopliteal lesions stenting with fasciotomy lamina vastoadductoria

**DOI:** 10.1101/2022.03.28.22272737

**Authors:** Alexey V. Cheban, Olesya S. Osipova, Pavel V. Ignatenko, Savr V. Bugurov, Alexandr A. Gostev, Shoraan B. Saaya, Artem A. Rabtsun, Andrey A. Karpenko

## Abstract

**Objective:** Fasciotomy can increase the mobility of the superficial femoral artery and reduce the incidence of stent breakage. This study aimed to compare the long-term patency of drug-eluting nitinol stents with and without fasciotomy in patients with prolonged SFA occlusions.

**Methods:** A randomized clinical trial was conducted in 60 (1:1) patients with long femoropopliteal steno-occlusive lesions more than 200 mm. Patients in group 1 (Zilver) underwent recanalization of occlusion of the femoropopliteal artery with stenting. In group 2 (ZilverFas), recanalization of the femoropopliteal occlusion with stenting and fasciotomy of Gunter’s canal were performed. The follow up evaluation of patency at 6, 12 months.

**Results:** 12-month primary patency in Zilver and ZilverFas groups was 51% and 80%, respectively (p = 0.02). The freedom from target revascularization (TLR) in Zilver and ZilverFas groups was 50% and 76%, respectively (p = 0.04). At one-years, primary-assisted and secondary patency for the ZilverFas and Zilver groups were 83% versus 62% (p = 0.07), 86% versus 65% (p = 0.05), respectively. In groups Zilver and ZilverFas, the number of stents fracture was 14 and 7, respectively (p = 0.05). The Cox multivariables regression indicated that the stent fracture, diabetes mellitus were the independent predictor of restenosis and reocclusion. Fasciotomy reduced the chance of reocclusion and restenosis by 2.94 times.

**Conclusions:** Our study has shown that a decompressing the stented segment with fasciotomy significantly improves the patency of the femoropopliteal segment and significantly reduces the number and severity of stents fractures.

## Introduction

Research on endovascular revascularization of long femoropopliteal lesions continues. Stent implantation in such lesions is less preferable, since the stent changes the biomechanics of the artery making it stiff. However, stent implantation is required in 5-15% of cases after balloon angioplasty due to dissection of the arterial intima [1]. Therefore, it is necessary to develop approaches that contribute to increasing the long-term patency of stents in the femoral-popliteal position. For example, drug-eluting stents have demonstrated better long-term patency compared to bare metal stents.

But any stent in the femoral-popliteal artery passing in the Gunter’s canal undergoes mechanical deformation (bending, twisting, stretching), which can lead to breakage of the stent [2]. Moreover, with an increase in the length of the implanted devices, the risk of fracture increases up to 60% due to the constantly changing configuration femoral artery [3]. Stent fracture can be a potential causative factor for stent restenosis.

To reduce the axial and physiological loads on the stented SFA we proposed a technique for increasing its functional mobility. The methods consist of the dissection the by Gunter’s canal fascia and crossing two pairs of arteries that bend around the knee joint. The safety of this procedure was confirmed by our pilot study [4].

This study aimed to compare the long-term patency of drug-eluting nitinol stents with and without fasciotomy in patients with prolonged SFA occlusions.

## Methods and materials

### Study design

A randomized clinical trial was conducted in patients with long femoropopliteal steno-occlusive lesions more than 200 mm (Fig. 1). This study was conducted following the principles of the Declaration of Helsinki and Guidelines for Good Clinical Practice. The ethics committee approved the study protocol and written informed consent was obtained from all patients before enrollment. The study is registered on ClinicaTrials.gov №NCT02590471.

**Figure 1:**
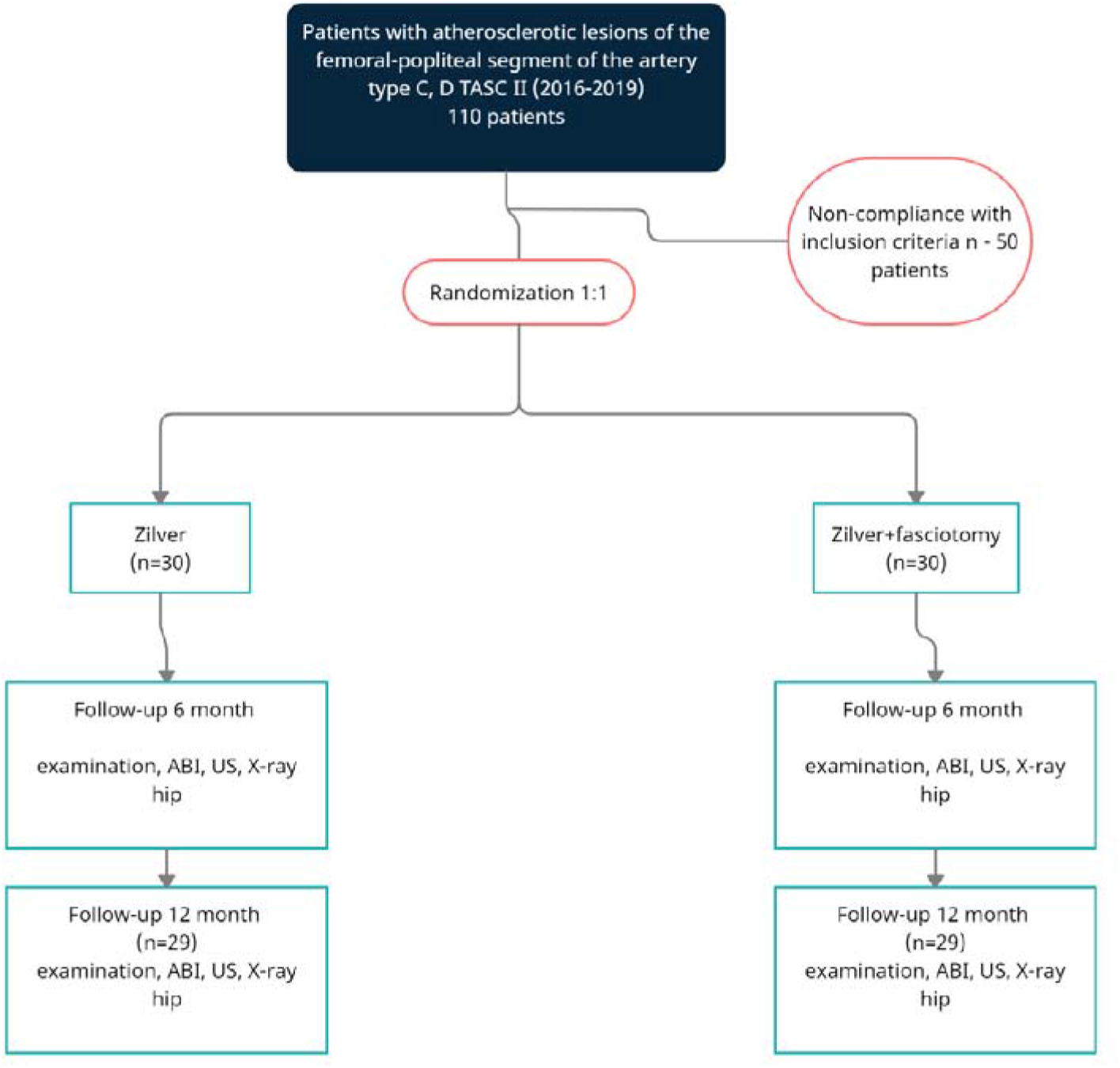
Study Design Flow Chart. Patient Disposition Flow Chart_CONSORT Diagram

### Sample size

It has been estimated that 60 patients should be included in the study to have a statistical power of 80% with alfa set at 5% (on a two-sided test) to detect an absolute difference in patency of 35% at 12 months. The absolute difference of 35% was selected based on the analysis of previous studies (assumed proportion of primary patency of 45% in Zilver group and 80% in ZilverFas group).

### Patients selection and data collection

The study was conducted in the period from 2016 to 2019. The included patients were 45 to 75 years of age, with chronic Rutherford 4-6 ischemia and satisfactory Rutherford outflow area. The inclusion and exclusion criteria are shown in Table 1. Thus, 60 patients with long lesions of femoropopliteal artery were included in the study. Randomization was done 1: 1 using the envelope method. All patients underwent a preoperative assessment using physical examinations, angiography or computed tomography angiography (CTA). Resistance values of runoff arteries were assessed using the Rutherford score. And the “blind bag” corresponds to the number 10, and the fully passable runoff is denoted by 1. Patients in group 1 (Zilver) underwent recanalization of occlusion of the femoropopliteal artery with stenting. In group 2 (ZilverFas), recanalization of the femoropopliteal occlusion with stenting and fasciotomy of Gunter’s canal were performed.

**Table 1:**
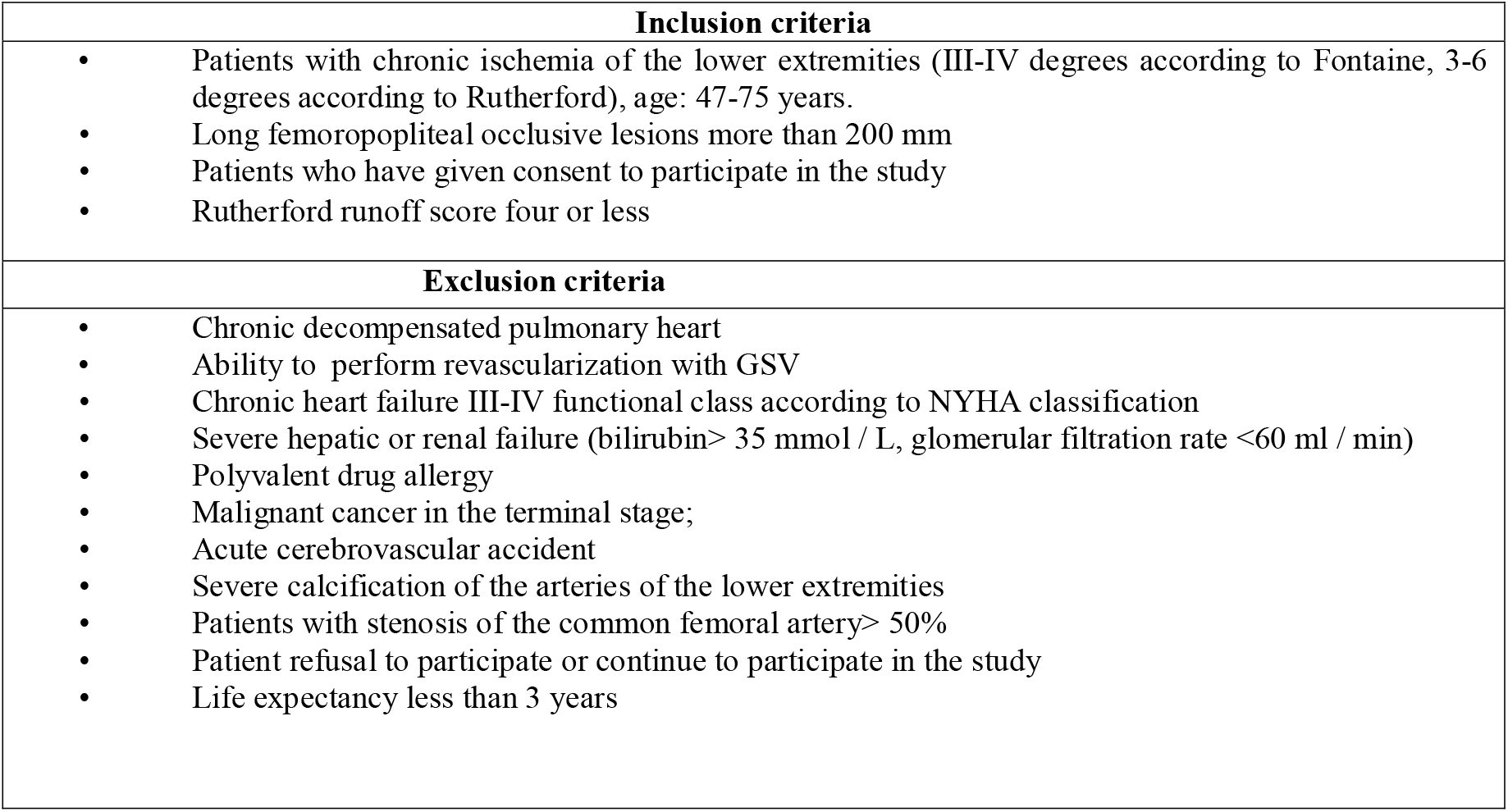
Inclusion and exclusion criteria

### Imaging

All CTA were obtained as part of routine clinical care. As per departmental policy, patients presenting with symptoms of claudication or chronic limb threatening ischemia undergo a duplex ultrasound first and then a CTA to plan intervention. All CTAs were performed using a protocol in helical scanning mode from the aortic bifurcation to the toes. Studies were collimated at 0.625 mm, with a peak kilovoltage of 120, automated milliamps and a rotation time of 64 0.75 s. Approximately 100 mL of iodinated contrast were administered at 4 mL/s using a power injector. Both duplex and CTAs were analyzed and reported as per the Society for Vascular Surgery reporting standards. All procedures were planned using the latest pre-operative CTA; duplex was used as the initial assessment method prior to CTA. During follow up (surveillance) all interventions were again planned using CTA, following an initial assessment with duplex.

### Interventions

Endovascular intervention was performed under local anesthesia with angiography visualization of the affected segment of the artery. During occlusion, transluminal or subintimal artery recanalization (most often mixed) was performed. Then balloon angioplasty of stenosis or occlusion was performed. After angiographic control, if necessary, a paclitaxel-eluting stent ZILVER PTX was placed along the entire length of the lesion.

In the ZilverFas group, under local anesthesia, the skin is dissected in the lower third of the thigh. Lamina Vastoadductoria was isolated, cut and tied, then the following arteries were cut and tied: a. superior medialis genus, a. superior lateralis genus. This technology is graphically presented in our article [5].

Drug therapy includes the appointment of aspirin (160-300 mg/day) and an intravenous heparin (5000 IU intravenously) before the procedure. After the procedure, all patients received aspirin at a dose of 100 mg/day for a long period, and clopidogrel at a dose of 75 mg/day for 3 months.

### Follow-up

The follow up evaluation of patency of the treated limb included symptoms assessment, physical examination, ankle-brachial index (ABI), duplex ultrasonography and a hip X-ray (to assess the stent fractures) at 6, 12 months. CTA was performed if restenosis more than 70% or occlusion was suspected.

### Definition

Technical success was defined as completion of the procedure and less than 30% residual stenosis. Primary patency was defined as absence of occlusion or flow-limiting stenosis (peak systolic velocity [PSV] ratio > 2.5 or stenosis more than 50% on duplex ultrasound/CTA) of the treated segment including 1 cm proximal and distal of the anastomosis. Primary assisted patency was defined as patency of the target lesion following re-intervention at the target vessel site in case of symptomatic restenosis. Secondary patency reports patency of the target lesion after treatment of a re-occlusion. Target lesion revascularization (TLR) was defined as a re-intervention at the index lesion. Clinical improvement was defined as change in Rutherford category, by at least 1 compared with baseline.

Major adverse cardiac events (MACE) were defined as myocardial infarction, stroke, or death. Major adverse limb events (MALE) defined as acute limb ischemia and major amputation of the index limb.

The severity of the stent fracture was determined according to the modified Rocha-Singh grading [6].

### Primary endpoint

12 month primary patency

### Secondary endpoints

technical success, intraoperative complications, limb salvage, 12 month secondary patency, stent fracture, wound complications.

### Statistical analysis

All data were analyzed on an intention-to-treat basis. We applied the Shapiro-Wilk W-test to verify the normality of the quantitative data distribution. Parametric variables were compared using Student t test. Nonparametric variables were compared using Manne Whitney U test. Categorical variables were compared using Fisher’s test. A log-rank test with a Kaplan-Meier curve was used to identify differences in the primary endpoint in the two groups. Cox proportional hazards models were used to determine the association of clinical, anatomical and procedural factors with loss patency. All p values <0.05 were considered statistically significant.

Statistical analyses of the study results was carried out using the STATISTICA 12.0 software («StatSoft», USA).

## Results

A total of 60 patients were enrolled, all of whom could be evaluated for the intention to treat analysis.

Each of the groups (Zilver and ZilverFas) included 30 patients with an average age of 63.3 ± 5.8 years and 65.7 ± 6.8 years, respectively.

### Baseline data

The clinical and lesions characteristics of the patients are presented in Table 2. No statistically significant differences were found. Stented lesion length in Zilver and ZolverFas groups was 220±22 mm and 220±17 mm.

**Table 2.** Baseline characteristics

| Characteristic | ZilverFas<br>(n=30) | Zilver<br>(n=30) | p |
| --- | --- | --- | --- |
| Gender, (male) n | 27 | 25 | 0,7 |
| Age, years | 65.7±6.8 | 63.3 ± 5.8 | 0.7 |
| Hospital Stay days | 7.7±2.1 | 4.7±2.1 | 0.01 |
| Current smoking, n (%) | 21 (70%) | 26 (87%) | 0.2 |
| Chronic heart failure, n (%) | 18 (60%) | 20 (67%) | 0.7 |
| Arterial hypertension, n (%) | 25 (83%) | 27 (90%) | 0.7 |
| Prior MI,n (%) | 6 (20%) | 8 (27%) | 0.76 |
| heart rhythm disturbance ,n (%) | 2 (17%) | 5 (17%) | 0.4 |
| Chronic kidney disease (%) | 16 (53%) | 23 (79%) | 0,1 |
| Hyperlipidemia (%) | 10 (38%) | 11 (41%) | 1 |
| Diabetes mellitus treatment, n (%) | 8 (27%) | 12 (49%) | 0.4 |
| ABI | 0.4±0.1 | 0.4±0.1 | 0.8 |
| Vessel runoff | 3.45 | 3.5 | 0.57 |
| Rutherford 4,n (%) | 22 (74%) | 26 (87%) | 0.3 |
| Rutherford 5,n (%) | 7 (23%) | 1 (3%) | 0.06 |
| Rutherford 6,n (%) | 1 (3%) | 3 (10%) | 0.6 |
| Lesion length, sm | 24.9±3.0 | 26±2.6 | 0.5 |
| Lesion type (occlusion, n (%)) | 30 (100%) | 30 (100%) | 0,07 |
| mean stent diameter, mm | 6,07 | 6,1 | 0,4 |
| Length of stented segment , mm | 220±17 | 220±22 | 1 |

### Main results

Postoperative complications were not statistically significant. Fasciotomy did not lead to an increase in the incidence of postoperative hematomas, lymphorrhea, and neuropathy, the patients in the groups did not have these complications. All wounds healed by primary intention.r Only a hospital stay in ZilverFas group was more longer. (Table 2)

12-month primary patency in Zilver and ZilverFas groups was 51% and 80%, respectively (p = 0.02). The data are presented in Fig. 2.

**Figure 2:**
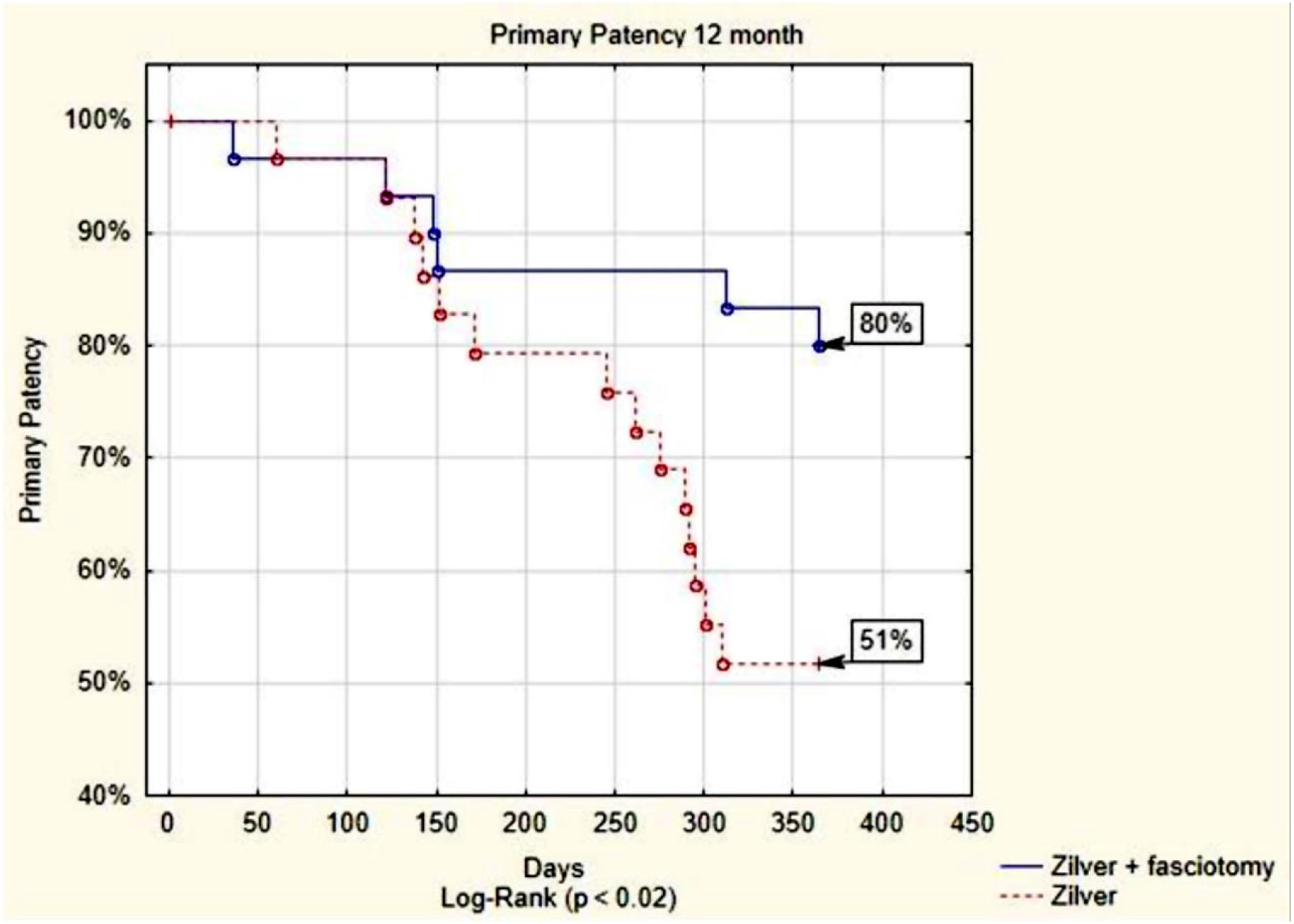
PPatency Kaplan-Meier Survival Curve

The freedom from target revascularization (TLR) in Zilver and ZilverFas groups was 50% and 76%, respectively (p = 0.04). In ZilverFas group, 7 patients underwent the re-intervention: three angioplasty with stenting, thre recanalizations with angioplasty and stenting, one femoral-popliteal bypass grafting. In Zilver group 15 patients underwent the re-intervention: nine angioplasty with stenting, five recanalizations with angioplasty and stenting, and one femoral-popliteal bypass.

At one-years, primary-assisted and secondary patency for the ZilverFas and Zilver groups were 83% versus 62% (p = 0.07), 86% versus 65% (p = 0.05), respectively. The long-term survival in Zilver group was 97%, and in ZilverFas group 94% (p=0.83).

In groups Zilver and ZilverFas, the number of stents fracture was 14 and 7, respectively (p = 0.05) (Fig. 3).

**Figure 3:**
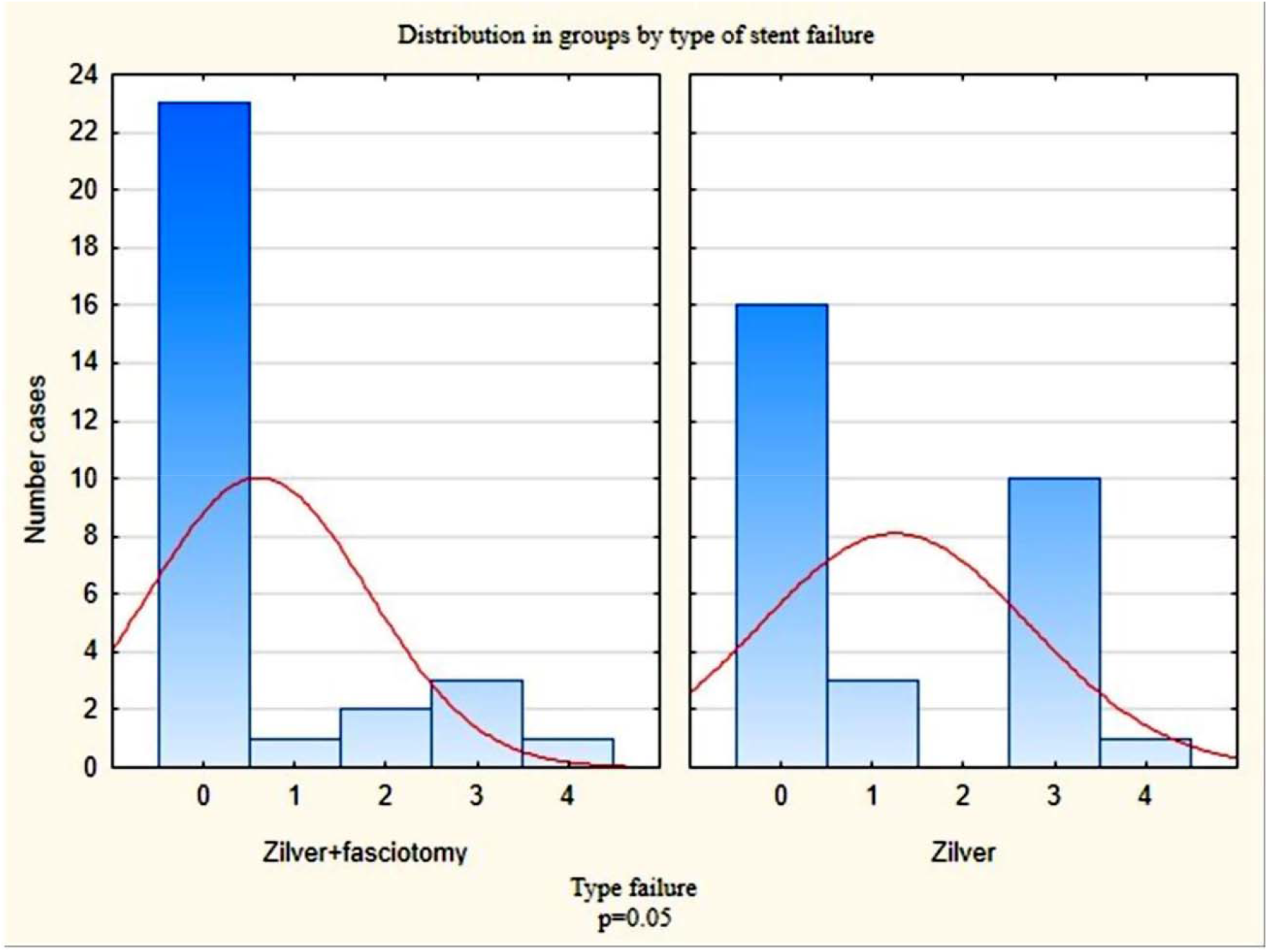
Distribution in groups by type of stent failure

The Cox multivariables regression indicated that the stent fracture, diabetes mellitus were the independent predictor of restenosis and reocclusion. Fasciotomy reduced the chance of reocclusion and restenosis by 2.94 times. (Table 3)

**Table 3.** Multivariable regression analysis factors associated with loss patency.

| Predictor | OR | p |
| --- | --- | --- |
| Residual stenosis | 1 [0,9; 1.02] | p 0.7 |
| Fracture | 2.8 [ 1.14; 6.7] | p 0.02 |
| Smoking | 0.8[0.29; 2.6] | p 0.8 |
| Coronary artery disease | 1.1[ 0.5; 2.6] | p 0.6 |
| Hypertension | 0.9[ 0.6; 1.33] | P 0.6 |
| Prior MI | 1.3[0.5; 3.39] | p 0,5 |
| Heart rhythm disturbance | 1.1[0.35; 4.09] | p 0,7 |
| Chronic heart failure | 0.6 [0.4; 1.08] | p 0,1 |
| Diabetes mellitus | 2.5[ 1.17; 5.67] | p 0.02 |
| Hyperlipidemia | 1.2 [ 0.5; 3.1] | p 0,5 |
| Male | 0.4[0.15;1.17] | p 0.1 |
| stenosis | 0.4 [ 0.1; 2.01] | p 0,2 |
| Occlusion | 2.1[0.49;9.22] | p 0.25 |
| Fasciotomy | 0,34 [0.13; 0.9] | P 0.02 |

During the 12 months MACE were 96.7% in each group. We didn’t have MALE in groups.

## Discussion

This randomized clinical trial compared one-year results of femoral-popliteal TASC C, D lesions stenting using the Zilver PTX drug-eluting stent with and without fasciotomy in 60 patients. Patients in the ZilverFas group showed a better 12-month primary patency compared to Zilver group. In our opinion, this was due to a significant decrease in the number of stent fractures after fasciotomy. Fasciotomy can reduce tension in the artery and increase its mobility.

During movement of the lower limb, the femoral-popliteal artery is subjected to various physiological stresses such as axial compression, radial compression, bending and twisting [7,8].

Moreover, the artery in the Gunter’s canal and behind the knee experiences the greatest load. These deformities are believed to play a major role in damage to the PBA wall and the impairment of blood flow [9]. Under the influence of such deformations, the rigid structure of the stent in the femoropopliteal artery undergoes high stress, which ultimately can lead to fracture of the stent [10,11].

Fractures of the stent in the SFA have previously been reported with an incidence of up to 65%, with 77% of stent fractures being the cause of restenosis [12]. Thanks to improved stent design, the breakdown rate was reduced to 3-12%, however, according to the literature, most studies were conducted in patients with short stenosis or occlusions, in some studies, there were patient losses at follow-up [3,13,14]. In our study, 20% of fractures in ZilverFas group and 50% in Zilver group were revealed, which can be explained by a high percentage of detectability because there were no losses on examinations and more severe damage to SFA.

In other studies, the annual primary patency after stenting of the femoropopliteal lesion TASS C, D is 40-50% [13,15,16]. Our primary patency results in Zilver group are consistent with these findings, but after fasciotomy, patency increased by 20%. In Zilver group, we observed twice as many stent fractures, and most of them were of grade 3 severity. Stent fractures increased the risk of target lesion loss of patency by 2.8 times. The same increase in risk has been shown in other studies[17,18].

Fasciotomy reduced the risk of restenosis or reocclusion by 2.94 times (OR=0.34), which confirms our hypothesis of decompression of the stented segment.

Diabetes was also an unfavorable predictor of loss of patency. Many studies have reported less success with endovascular treatment in diabetic patients[19,20,21].

## Conclusion

Our study has shown that a modified approach by decompressing the stented segment in the gunter’s canal with ligation of a.genus superior lateralis and medialis significantly improves the patency of the femoropopliteal segment and significantly reduces the number and severity of fractures. This technology is of interest for further research with various types of stents.

## Data Availability

All data produced in the present work are contained in the manuscript

